# Evaluation and modelling of the performance of an automated SARS-CoV-2 antigen assay according to sample type, target population and epidemic trends

**DOI:** 10.1101/2022.01.14.22269064

**Authors:** Nicolas Yin, Cyril Debuysschere, Valery Daubie, Marc Hildebrand, Charlotte Martin, Sonja Curac, Fanny Ponthieux, Marie-Christine Payen, Olivier Vandenberg, Marie Hallin

**Affiliations:** Department of Microbiology, Laboratoire Hospitalier Universitaire de Bruxelles – Universitair Laboratorium Brussel (LHUB-ULB), Université Libre de Bruxelles (ULB), Brussels, Belgium; Department of Internal Medicine, Erasme University Hospital, Université Libre de Bruxelles (ULB), Brussels, Belgium; Department of Infectious Diseases, Saint-Pierre University Hospital, Université Libre de Bruxelles (ULB), Brussels, Belgium; Emergency Department, Erasme University Hospital, Université Libre de Bruxelles (ULB), Brussels, Belgium; Centre for Environmental Health and Occupational Health, School of Public Health, Université Libre de Bruxelles (ULB), Brussels, Belgium; Clinical Research and Innovation Unit, Laboratoire Hospitalier Universitaire de Bruxelles – Universitair Laboratorium Brussel (LHUB-ULB), Université Libre de Bruxelles (ULB), Brussels, Belgium; Division of Infection and Immunity, Faculty of Medical Sciences, University College London, London, United Kingdom

**Keywords:** SARS-CoV-2, COVID-19, model, diagnostic, test, assay

## Abstract

The Lumipulse® G SARS-CoV-2 Ag assay performance was evaluated on prospectively collected saliva and nasopharyngeal swabs (NPS) of recently ill in- and outpatients and according to the estimated viral load. Performances were calculated using RT-PCR positive NPS from patients with symptoms ≤ 7 days and RT-PCR negative NPS as gold standard. In addition, non-selected positive NPS were analyzed to assess the performances on various viral loads. This assay yielded a sensitivity of 93.1% on NPS and 71.4% on saliva for recently ill patients. For NPS with a viral load > 103 RNA copies/mL, sensitivity was 96.4%. A model established on our daily routine showed fluctuations of the performances depending on the epidemic trends but an overall good negative predictive value. Lumipulse® G SARS-CoV-2 assay yielded good performance for an automated antigen detection assay on NPS. Using it for the detection of recently ill patient or to screen high-risk patients could be an interesting alternative to the more expensive RT-PCR.

## 1. Introduction

The Lumipulse® G SARS-CoV-2 Ag assay is an automated assay allowing SARS-CoV-2 antigen quantification on universal transport medium (UTM) preserved nasopharyngeal swabs (NPS) but also saliva. Its performance was assessed previously [1,2]. However, data are missing regarding recently symptomatic patients (≤ 7 days since symptom onset (DSO)) and the use of saliva samples. Saliva sample use could indeed improve the comfort of patients, and decrease the needs in terms of swabs and transport media [3]. Furthermore, existing studies did not evaluate the pretreatment step recommended by the manufacturer for viral deactivation and sample fluidification. Adding a deactivation step does improve biosafety and technical failure rate, but can also decrease the sensitivity of the test [4]. Here, we evaluated the performance of the Lumipulse® G SARS-CoV-2 assay using the manufacturer’s sample extraction solution (SES) pretreatment on both NPS and saliva and assessed its performance with regards to the target population and the viral load. In a second time, we extrapolated its results on the daily routine of our laboratory in a model assessing the potential influence of epidemic trends on its overall performances.

## 2. Materials and Methods

### 2.1. Study design, population and sample collection

A minimum of 500 residual nasopharyngeal swabs preserved in 3 mL UTM from patients with a SARS-CoV-2 RT-PCR performed on a routine basis in our laboratory were selected. The selection included a minimum of 300 SARS-CoV-2 negative RT-PCR samples from outpatients, 100 SARS-CoV-2 negative RT-PCR samples from inpatients and 100 SARS-CoV-2 positive RT-PCR samples from patients with maximum 7 DSO. In addition, residuals of non-selected positive samples with either more than 7 DSO or an unknown date of symptoms were added to assess the limit of detection. All these samples were stored at 4°C and analyzed less than 2 days after the RT-PCR was performed.

Saliva samples were collected from patients prospectively enrolled in two settings: outpatients at the consultation or at the sampling center with a prescription of a SARS-CoV-2 PCR test on NPS as well as inpatients with a positive SARS-CoV-2 RT-PCR test on NPS for less than 2 days.

### 2.2. SARS-CoV-2 RT-PCR

SARS-CoV-2 RT-PCR on nasopharyngeal swab was considered as the gold-standard. If the nasopharyngeal swab was not available anymore to perform the assay, the RT-PCR performed on saliva was considered as a proxy of the gold standard.

RT-PCR were performed using the Alinity m SARS-CoV-2 assay (Abbott Molecular, USA). The two target sequences are located in the RdRp and N genes of the SARS-CoV-2 genome and are detected using the same fluorophore. Saliva samples were diluted before extraction four times to reach the volume of 500μL needed for this assay. Cycle threshold (Ct) values were plotted with standards provided by the Belgian national reference center following recommendations by Sciensano to provide semi-quantitative results [5]. The correspondence table between Ct values and viral load is summarized in table 1.

**Table 1.** Semi-quantification of SARS-CoV-2 RT-PCR results using the Alinity m SARS-CoV-2 assay (Abbott Molecular, USA).

| Semi-quantification | Ct values | Viral load (RNA copies /mL) |
| --- | --- | --- |
| Weak | > 29.9 | < 10<sup>3</sup> |
| Mild | > 23.3 - 29.9 | 10<sup>3</sup> - < 10<sup>5</sup> |
| Strong | > 16.7 - 23.3 | 10<sup>5</sup> - < 10<sup>7</sup> |
| Very strong | ≤ 16.7 | ≥ 10<sup>7</sup> |

### 2.3. SARS-CoV-2 antigen quantification

Antigen quantification (Ag) was performed using the Lumipulse® G SARS-CoV-2 Ag assay, expressing the dosage in pg/mL. Viral deactivation was performed using the sample extraction solution (SES) by diluting 4 part of UTM with 1 part of SES or 1 part of saliva with 1 part of SES. A 30 minutes incubation at room temperature was performed before centrifugation at 2000g for 5 minutes. Lumipulse SARS-CoV-2 antigen testing was performed on a Lumipulse® G600II instrument according to manufacturer’s instruction. Positivity threshold was determined using the positive samples from patients with ≤ 7 DSO and all the negative samples using the following criteria: highest possible Youden’s index, specificity > 98% and a minimum of 0.6 pg/mL (instrument quantification limit)

### 2.4. SARS-CoV-2 variant determination

Samples with a minimal SARS-CoV-2 viral load of 105 RNA copies/mL were randomly sequenced for surveillance purpose.

### 2.5. Model

Positive RT-PCR results performed routinely in our laboratory are semi-quantified according to their Ct values regardless of their indication (symptoms, screening…). Daily overall sensitivity (Se), positive predictive value (PPV) and negative predictive value (NPV) were estimated according to the performance evaluation results by semi-quantification level. To minimize day-to-day and holiday-related fluctuations, data were computed from May 1, 2020 to October 30, 2021 using a backward sliding window of 14 days (hereafter referred as “14-day Se”, “14-day PPV” and “14-day NPV”).

### 2.6. Statistical analyses

Statistical analyses and receiver operating characteristic (ROC) curves were performed using Analyse-it® for Microsoft Excel v5.30.4.

### 2.7. Ethical approval

Erasme university hospital ethics’ committee approved and reviewed the comparative performance evaluation study on saliva and nasopharyngeal swabs. Ethics’ comitee of Saint-Pierre hospital waived ethical approval for the use of residual human body material for evaluation purpose.

## 3. Results

### 3.1. Population

The studied samples are summarized in Table 2.

**Table 2.** Study population (N: total number of samples, Ag: antigen quantification, NPS: nasopharyngeal swabs, DSO: days since symptom onset).

| Samples collected | N | Ag (NPS) | Ag (Saliva) |
| --- | --- | --- | --- |
| Overall | 632 | 605 | 144 |
| Negative RT-PCR | 408 | 400 | 83 |
| Outpatients | 304 | 300 | 76 |
| Inpatients | 104 | 100 | 7 |
| Positive RT-PCR | 224 | 205 | 61 |
| $\leq 7$ DSO | 116 | 102 | 35 |
| Alpha variants | 61 | 61 | 14 |
| Gamma variants | 8 | 8 | 2 |
| Delta variants | 22 | 22 | 0 |
| Not a variant of concern | 2 | 2 | 1 |

### 3.2. Threshold determination

#### 3.2.1. NPS

Five hundred and two samples were selected including 102 positive samples with ≤ 7 DSO. ROC curve analysis yielded an area under the curve (AUC) at 0.973±0.023 (Figure 1). The highest Youden Index was at a threshold of 2.47 pg/mL (sensitivity 93.1%, specificity 99.0%). Analytical performance and their confidence interval at this threshold are summarized in table 3.

**Figure 1.**
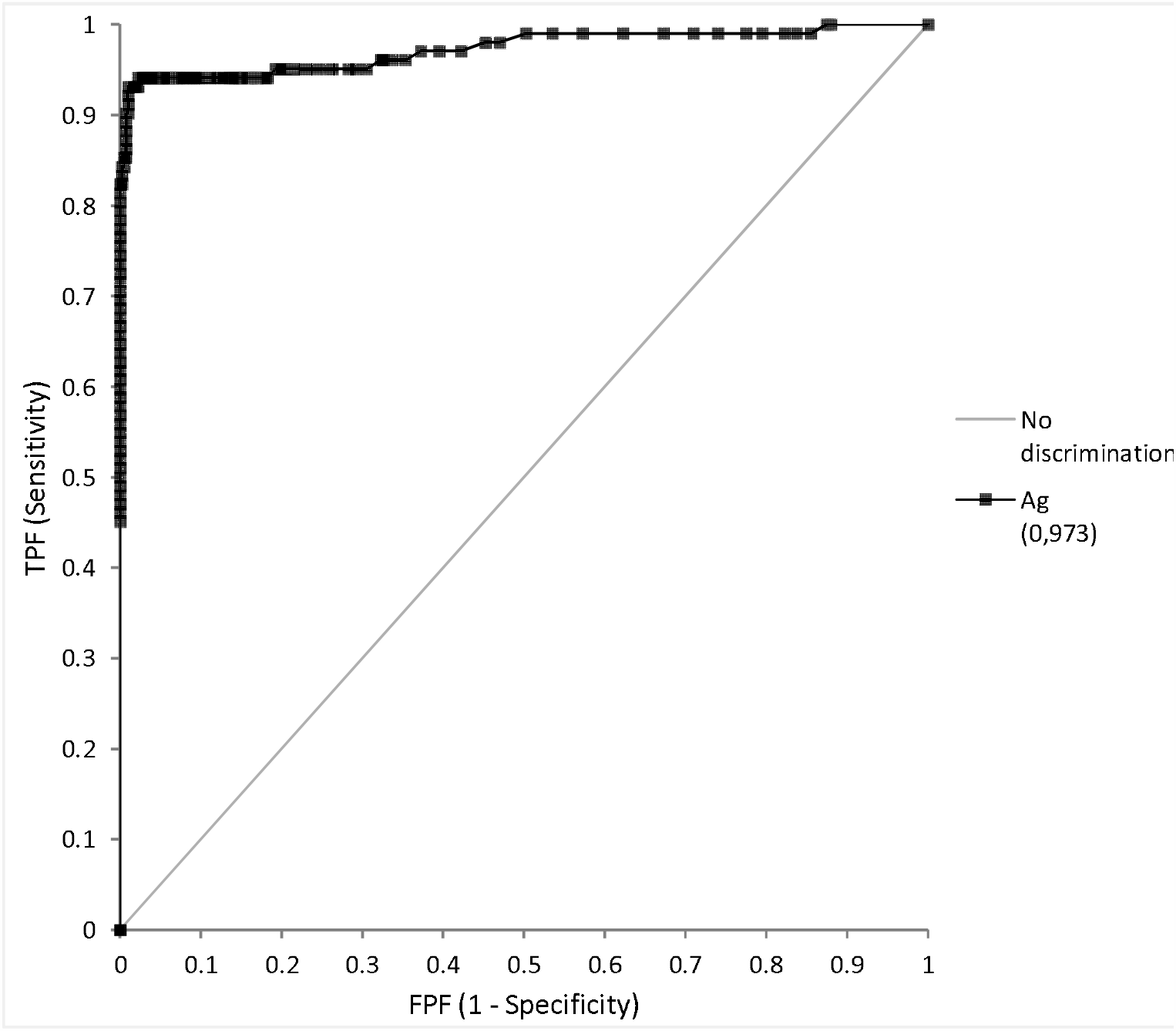
Receiver Operating Curve (ROC) analysis of antigen quantification on nasopharyngeal swabs (NPS) vs RT-PCR on NPS as Gold standard.

**Table 3.** Analytical performance of the Lumipulse G SARS-CoV-2 assay on nasopharyngeal swabs (NPS) and saliva vs RT-PCR on NPS. T: positivity threshold, CI: confidence interval, DSO: days since symptom onset.

| Samples collected | Ag (NPS, T = 2.47 pg/mL) |  | Ag (Saliva, T = 0.60 pg/mL) |  |
| --- | --- | --- | --- | --- |
|  | N | % (Wilson 95% CI) | N | % (Wilson 95% CI) |
| Sensitivity (DSO $\leq 7$) | 95/102 | 93.1 (86.5-96.6) | 25/35 | 71.4 (54.9-83.7) |
| Specificity (overall) | 396/400 | 99.0 (97.5-99.6) | 82/83 | 98.8 (93.5-99.8) |
| Outpatients | 298/300 | 99.3 (97.6-99.8) | 75/76 | 98.7 (92.9-99.8) |
| Inpatients | 98/100 | 98.0 (93.0-99.4) | 7/7 | 100 (64.6-100) |

#### 3.2.2. Saliva

One hundred eighteen samples were selected including 35 positive samples with ≤ 7 DSO. ROC curve analysis yielded an area under the curve (AUC) at 0.973±0.023 (Figure 2). The highest Youden Index was at a threshold of 0.55 pg/mL (sensitivity 71.4%, specificity 98.8%). To respect the quantification limit, the positivity threshold was set at 0.6 pg/mL. Analytical performance and their confidence interval at this threshold are summarized in table 3.

**Figure 2.**
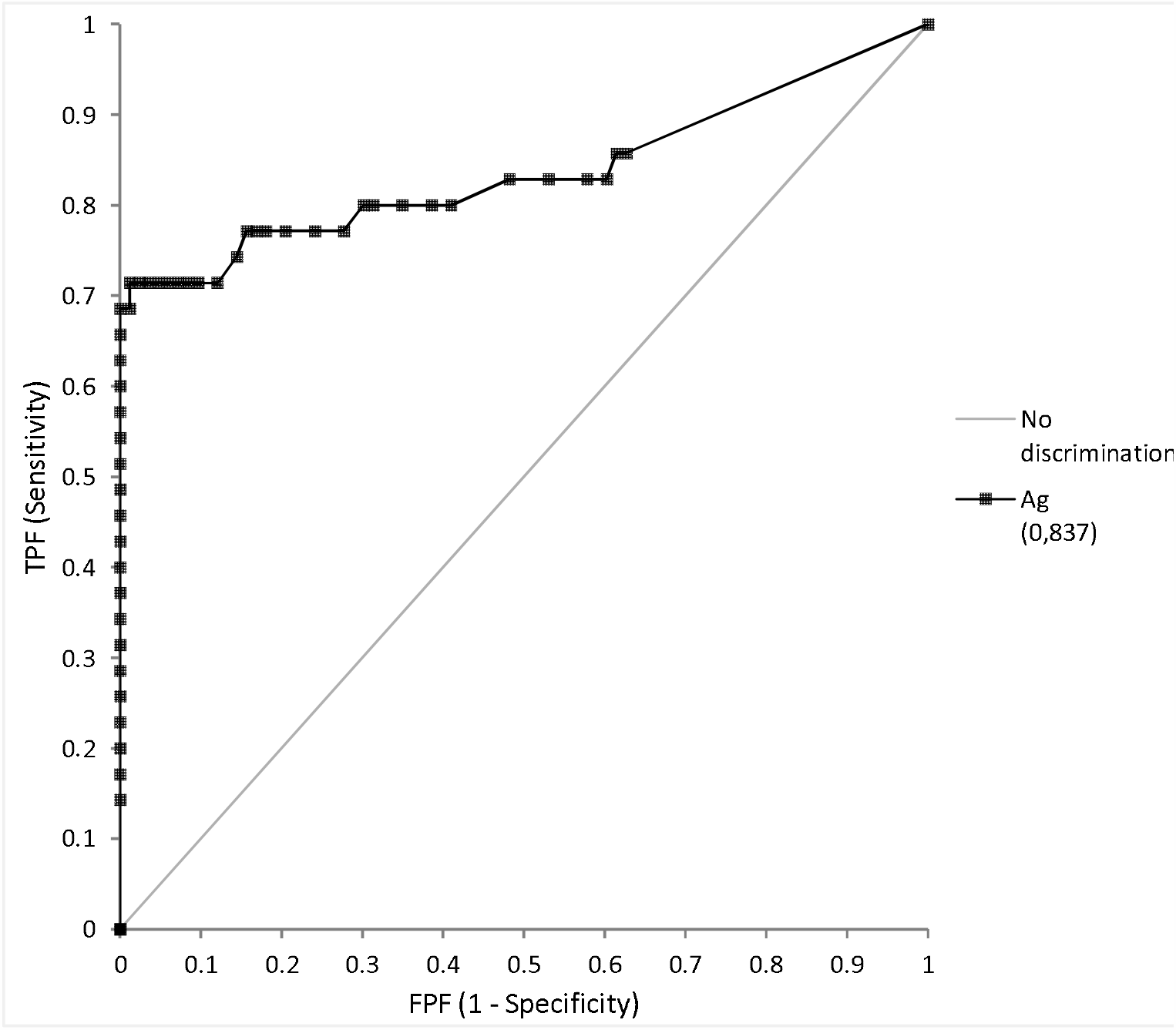
Receiver Operating Curve (ROC) analysis of antigen quantification on saliva vs RT-PCR on nasopharyngeal swabs (NPS) as Gold standard.

### 3.3. Detection limit assessment

Sensitivity of antigen quantification was assessed at 4 different levels of RT-PCR semi-quantification on NPS and are summarized in table 4.

**Table 4.** Analytical performance of the Lumipulse G SARS-CoV-2 assay on nasopharyngeal swabs (NPS) and saliva vs RT-PCR on NPS at different level of viral load (T: positivity threshold, CI: confidence interval, Ct: cycle threshold value).

| Viral load (RNA copies /mL) | Ag (NPS, T = 2.47 pg/mL) |  | Ag (Saliva, T = 0.60 pg/mL) |  |
| --- | --- | --- | --- | --- |
|  | N | % (Wilson 95% CI) | N | % (Wilson 95% CI) |
| $< 10^3$ (Ct $> 29.9$) | 4/38 | 10.5 (4.2-24.1) | 5/8 | 62.5 (30.6-86.3) |
| $\geq 10^3$ (Ct $\leq 29.9$) | 141/167 | 96.4 (92.4-98.3) | 32/46 | 69.6 (55.2-80.9) |
| $10^3 - <10^5$ | 36/41 | 87.8 (74.5-94.7) | 8/19 | 42.1 (23.1-63.7) |
| $10^5 - <10^7$ | 70/71 | 98.6 (92.4-99.8) | 12/15 | 80.0 (54.8-93.0) |
| $\geq 10^7$ | 55/55 | 100 (93.5-100) | 12/12 | 100 (75.8-100) |

### 3.4. Variants of interest

All the successfully sequenced positive samples yielded a positive antigen result on NPS at the previously determined threshold; no variant effect was highlighted.

### 3.5. Modelling the influence of epidemic trends on the overall performance of the assay

The model showed that using the automated SARS-CoV-2 antigen quantification on our day-to-day non-selected routine samples would exhibit important variation of sensitivity and predictive positive value according to the dynamic of the epidemic at the time (Figure 3). Indeed, during low circulation phases, sensitivity decreases sharply due to a very high proportion of weak positive samples. Likewise, a higher proportion of false positive penalizes the positive predictive value. Conversely, during epidemic peaks, the positive predictive value increased at more than 90% in November, 2020 while the negative predictive value decreased slightly below 95% like after the end of the first epidemic wave in Belgium in May, 2020.

**Figure 3.**
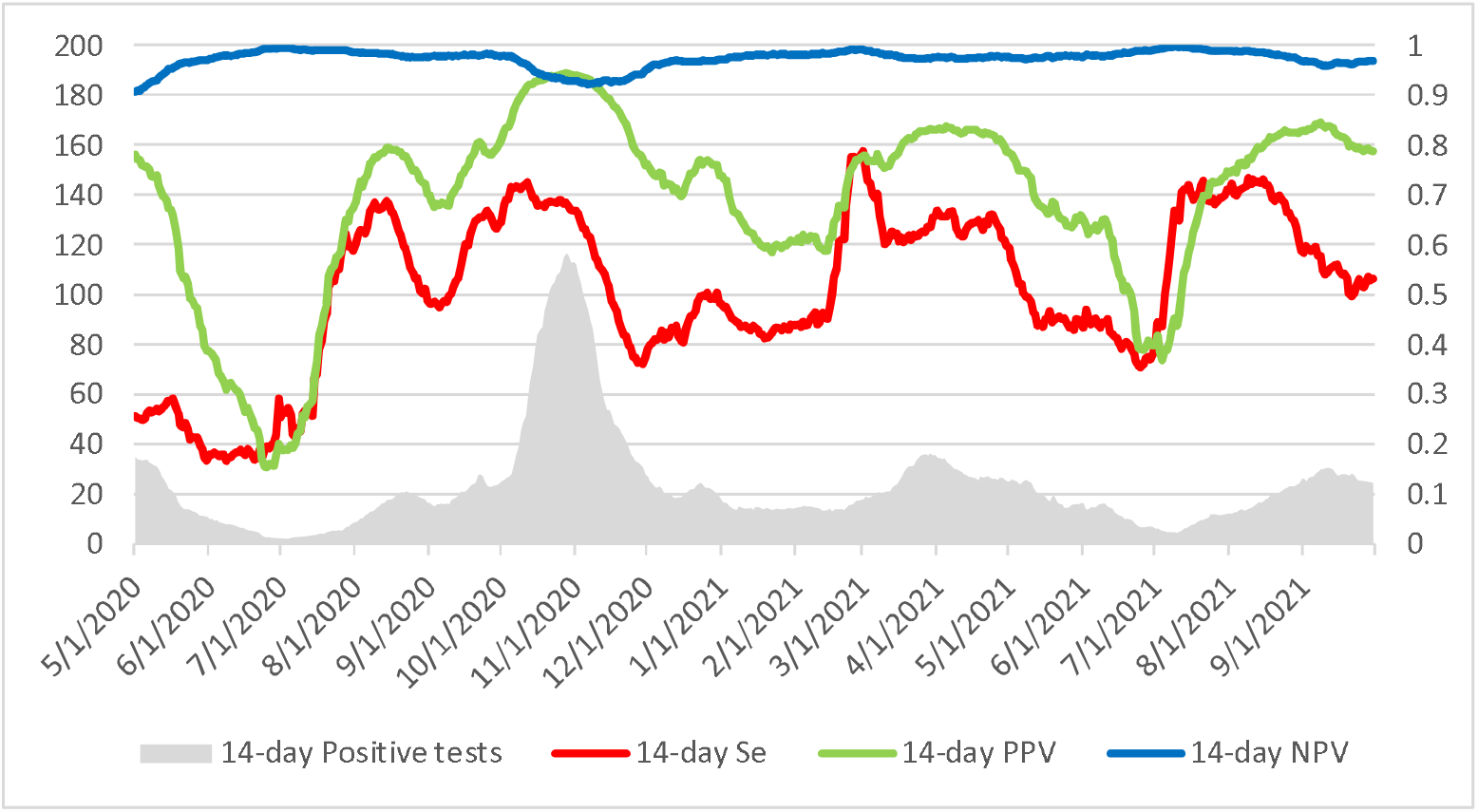
Modelling SARS-CoV-2 automated antigen detection performance on epidemic trends. Data computed from May 1, 2020 to October 30, 2021 using a backward sliding window of 14 days (14-day Se: 14-day sensitivity, 14-day PPV: 14-day Positive Predictive Value, 14-day NPV: 14-day Negative Predictive Value).

## 4. Discussion

In a previous study, we proposed to position the different available assays for the direct diagnostic of COVID-19 in a structured algorithm. Rapid antigen tests were dedicated to the fast diagnostic of symptomatic patients and automated antigen quantification assays were used as a screening method. This strategy allows providing a faster and cheaper but reliable alternative to large molecular platforms [2]. In the present study, the Lumipulse® G SARS-CoV-2 assay yielded excellent analytical performance on NPS for recently symptomatic patients with a sensitivity of 93.1% for patients with DSO ≤ 7 for a specificity of 99.0%. Performances have improved compared to our previous study thanks to the use of the manufacturer deactivation method. Performances of the Lumipulse® G SARS-CoV-2 on NPS were even closer to those of RT-PCR when excluding low positive samples (Se = 96.4%), demonstrating its particular interest to (i) diagnose recently ill patient and (ii) efficiently detect infectious people in a wider screening strategy. As expected, saliva yielded less good performance and should be restricted to settings with limited access to sampling disposable or patients presenting contra-indication for naso-pharyngeal sampling. Comparable results were previously obtained on saliva and on other instruments [6–8].

Additionally, we modelled the estimated performance of this automated quantification assay on our day-to-day routine (“all comers” non-selected samples). As expected, our laboratory being a hospital laboratory, an important proportion (46.8%) of samples were weak positive samples, which impaired in this model the overall sensitivity and positive predictive value of the test as compared with results obtained during the analytical evaluation. However, using this extrapolation could be of use to choose the best technique on the best target population in their own setting. Despite this non-selection of samples, the negative predictive value stayed over 95% during the epidemic peaks. This good negative predictive value and the good sensitivity on samples with significant viral load confirm that the use of this technique is interesting for the fast and large screening of asymptomatic patients in contexts such as hospital admission, participation of a social event and travel. Indeed, this technique was implemented at several airports in Germany to screen travelers using oropharyngeal swabs [9], and in Italy in schools using saliva [10]. However, during periods of low virus circulation, this technique should be used in a 2-step strategy, with positive samples checked by a reflex RT-PCR, to balance the lower positive predictive value observed during these periods. From a public health perspective, such strategy would decrease dramatically the costs of the screening for a limited risk and free the PCR instruments for more serious cases and other uses.

Furthermore, automation allows a higher throughput than manual antigen rapid testing (60-100 tests per hour per instrument) and provides an automated, objective reading of the test. Furthermore, the use of UTM also allows secondary PCR checks and sequencing on positive samples.

Our results provide evidence that the Lumipulse® G SARS-CoV-2 Ag assay is a robust antigen quantification assay for the detection of SARS-CoV-2 on UTM preserved NPS especially for recently ill patients and people with high viral load. The use of this technique could offer a fast and cost-efficient solution for diverse situations such as systematic or targeted screening in specific situations (travelers, social events, nightlife…).

## Data Availability

All data produced in the present work are contained in the supplementary material.

## Supplementary Materials

The following supporting information can be downloaded: Table S1: Dataset.

## Author Contributions

Conceptualization, Nicolas Yin, Olivier Vandenberg and Marie Hallin; Data curation, Nicolas Yin; Formal analysis, Nicolas Yin; Funding acquisition, Olivier Vandenberg; Investigation, Nicolas Yin, Cyril Debuysschere, Valery Daubie, Marc Hildebrand, Charlotte Martin, Sonja Curac, Fanny Ponthieux and Marie-Christine Payen; Methodology, Nicolas Yin; Resources, Olivier Vandenberg and Marie Hallin; Supervision, Marie Hallin; Validation, Nicolas Yin, Olivier Vandenberg and Marie Hallin; Writing – original draft, Nicolas Yin; Writing – review & editing, Olivier Vandenberg and Marie Hallin..

## Funding

This research was funded by Fujirebio. The authors did not perceive any personal grants or funding.

## Institutional Review Board Statement

The study was conducted in accordance with the Declaration of Helsinki, and approved by the Ethics Committee of Erasme Hospital (protocol code P2021/104/B4062021000050, approved 2 March 2021).

## Informed Consent Statement

Informed consent was obtained from all subjects involved in the performance evaluation study on saliva.

No written consent was required for the use of residual human body material for evaluation purpose.

## Data Availability Statement

The data presented in this study are available in supplementary material.

## Acknowledgments

We wish to thank the personnel of the LHUB-ULB for its daily technical assistance. This work is dedicated to the healthcare workers, the patients, and families affected by SARS-CoV-2.

## Conflicts of Interest

The authors declare no conflict of interest. The funders had no role in the design of the study; in the collection, analyses, or interpretation of data; in the writing of the manuscript, or in the decision to publish the results.

## Notes

### Competing Interest Statement

The authors have declared no competing interest.

### Author Declarations

Erasme university hospital ethics' committee approved and reviewed the comparative performance evaluation study on saliva and nasopharyngeal swabs. Ethics' comittee of Saint-Pierre hospital waived ethical approval for the use of residual human body material for evaluation purpose.

